# Genome Wide Epistasis Study of On-Statin Cardiovascular Events with Iterative Feature Reduction and Selection

**DOI:** 10.1101/2020.03.31.20044255

**Authors:** Solomon M. Adams, Habiba Feroze, Tara Nguyen, Seenae Eum, Cyrille K. Cornelio, Arthur F. Harralson

**Author notes:** **Corresponding Author:** Solomon M. Adams, PharmD, PhD, Shenandoah University School of Pharmacy, 8095 Innovation Park Drive, Suite 301, Fairfax, VA 22031-4868. HF and TN contributed equally.

## Abstract

**Background:** Predicting risk for major adverse cardiovascular events (MACE) is an evidence-based practice that incorporates lifestyle, history, and other risk factors. Statins reduce risk for MACE by decreasing lipids, but it is difficult to stratify risk following initiation of a statin. Genetic risk determinants for on-statin MACE are low-effect size and impossible to generalize. Our objective was to determine high-level epistatic risk factors for on-statin MACE with GWAS-scale data.

**Methods:** Controlled-access data for 5,980 subjects taking a statin collected from Vanderbilt University Medical Center’s BioVU were obtained from dbGaP. We used Random Forest Iterative Feature Reduction and Selection (RF-IFRS) to select highly informative genetic and environmental features from a GWAS-scale dataset of patients taking statin medications. Variant-pairs were distilled into overlapping networks and assembled into individual decision trees to provide an interpretable set of variants and associated risk.

**Results:** 1,718 cases who suffered MACE and 4,172 controls were obtained from dbGaP. Pathway analysis showed that variants in genes related to vasculogenesis (FDR=0.024), angiogenesis (FDR=0.019), and carotid artery disease (FDR=0.034) were related to risk for on-statin MACE. We identified six gene-variant networks that predicted odds of on-statin MACE. The most elevated risk was found in a small subset of patients carrying variants in *COL4A2, TMEM178B, SZT2*, and *TBXAS1* (OR=4.53, p<0.001).

**Conclusions:** The RF-IFRS method is a viable method for interpreting complex “black-box” findings from machine-learning. In this study, it identified epistatic networks that could be applied to risk estimation for on-statin MACE. Further study will seek to replicate these findings in other populations.

## Introduction

Predicting risk for Cardiovascular Disease (CVD) is a mainstay of primary care and cardiology. Patients who develop CVD are at risk for major adverse cardiovascular events (MACE), such as myocardial infarction, stroke, or unstable angina. Risk assessments for CVD include clinical biomarkers, family history, lifestyle, co-morbidities and biometrics. Routine risk assessments for CVD risk guide major therapeutic and lifestyle decisions.

Hyperlipidemia is a risk factor for CVD and MACE, and the American College of Cardiology (ACC) guidelines on the management of blood cholesterol recommend statins as the cornerstone pharmacotherapy.^1^ CVD risk reduction from statins might be population specific and shows diversity among different patient groups. Ramos and colleagues found that the incidence of MACE was 19.7 (statin-users) and 24.7 (statin non-users) events per 1,000 person-years in patients with asymptomatic peripheral artery disease.^2^ Another study concluded that statin therapy had no major benefit on stroke in women.^3^ Overall, however, statins reduce the risk for MACE proportional to the magnitude of cholesterol lowering in all ages.^4^

Pharmacogenomics (PGx) guidance supports PGx for prevention of myopathy with simvastatin based on patients’ *SLCO1B1* haplotype.^5^ Additionally, statin biochemical response (e.g. PK, Lipid Lowering Efficacy) is associated with numerous variations. Ruiz-Iruela and colleagues found that decreased lipid lowering of rosuvastatin, atorvastatin, and simvastatin is predicted by *ABCA1* rs2230806 and *CYP2D6* *3. They also found that *CETP* variants rs708272 and rs5882 were associated with decreased and increased LDL lowering with rosuvastatin, respectively.^6^ These variations, however, have not been found to be associated with higher level outcomes like prevention of CVD-related events. Low-effect size risk variants also provide insight into pathogenesis of CVD. Genetic variations in apolipoprotein C-III (*APOC3*) and angiopoietin-like 4 (*ANGPTL4*) have been associated with risk for coronary artery disease (CAD).^7^ Roguin and colleagues found that the Haptoglobin (*HP*) genotype was a significant independent predictor of MACE in patients with diabetes.^8^ The PROSPER study found that *SURF6* rs579459 was associated with CAD, stroke and large artery stroke. It also found that *TWIST1* rs2107595 was associated with an increased risk of MACE such as large artery stroke, CAD, and ischemic stroke.^9^

Routine genetic testing for hyperlipidemia and CVD risk is limited to patients with history of familial hypercholesterolemia (FH), predicted by variation in *LDLR, APOB*, or *PCSK9*.^1^ CVD nevertheless shows strong heritability in patients without FH, suggesting an underlying genetic component.^7^ Genome-Wide Association Studies (GWAS) have identified over 50 genetic variants that are associated with risk for CVD and MACE. Clinical translation of these genetic risk factors is challenged by individual variants with small effect sizes and poor understanding of the interplay between multiple genetic variants and risk for MACE. These genetic factors might help explain cases in which patients still experience MACE in spite of adequate phamacologic response to statin therapy and other risk reduction strategies.

PGx is exemplified in variations among drug-metabolizing genes, including phase I (oxidation, reduction, hydrolysis), phase II (conjugation), and phase III (transport). In these cases, functional genetic variations can have catastrophic effects on pharmacokinetics.^10^ While some evidence supports PGx for pharmacodynamic markers, PGx outside of pharmacokinetics has been limited by relatively low effect-size of individual variants, and the inability to consistently apply multiple gene effects. This is partially addressed by the growing use of polygenic risk scores (PRS) to pool effects from unrelated variants;^11^ however, little has been done to incorporate the effects of epistasis (i.e. gene-gene interactions) to create novel predictors of drug response.

The objective of this research was to stratify the risk of on-statin MACE based on polygenic epistatic predictors. We applied a step-wise, interpretable, machine-learning (ML) driven ensemble method for feature reduction and determination of epistasis to a GWAS-scale dataset. We expect that application of this method will drive novel insight into genetic interactions that drive risk for complex cardiovascular phenotypes and statin PGx.

## Methods

### Clinical Dataset

The data/analyses presented in the current publication are based on the use of controlled-access study data downloaded with permission from the dbGaP web site, under phs000963.v1.p1 (https://www.ncbi.nlm.nih.gov/projects/gap/cgi-bin/study.cgi?study_id=phs000963.v1.p1). This dataset was assembled through Vanderbilt University Medical Center’s BioVU repository and clinical data was extracted from the electronic medical record. The study was approved by the Vanderbilt institutional review board and all subjects provided informed consent. These data correspond to 5,890 subjects of European descent taking HMG-Coa Reductase Inhibitors (statins) who were genotyped with the Illumina HumanOmniExpressExome 8v1-2_A array by the RIKEN Integrative Medical Sciences Center (IMS) and supported by the Pharmacogenomics Research Network (PGRN)-RIKEN IMS Global Alliance. Inclusion and exclusion criteria for cases and controls is described in **Table <u>1</u>**. The primary outcome is on-statin MACE, defined as any revascularization event (e.g. stent placement, bypass) and/or acute myocardial infarction. The case group contains 1,718 subjects, and the control group contains 4,172 subjects. Case and control status was determined with Vanderbilt’s BioVU DNA databank and associated Synthetic Derivative database of clinical information, and software tools developed to identify drugs and clinical events using Electronic Health Record-derived structured and unstructured (“free text”) data.

**Table 1:**
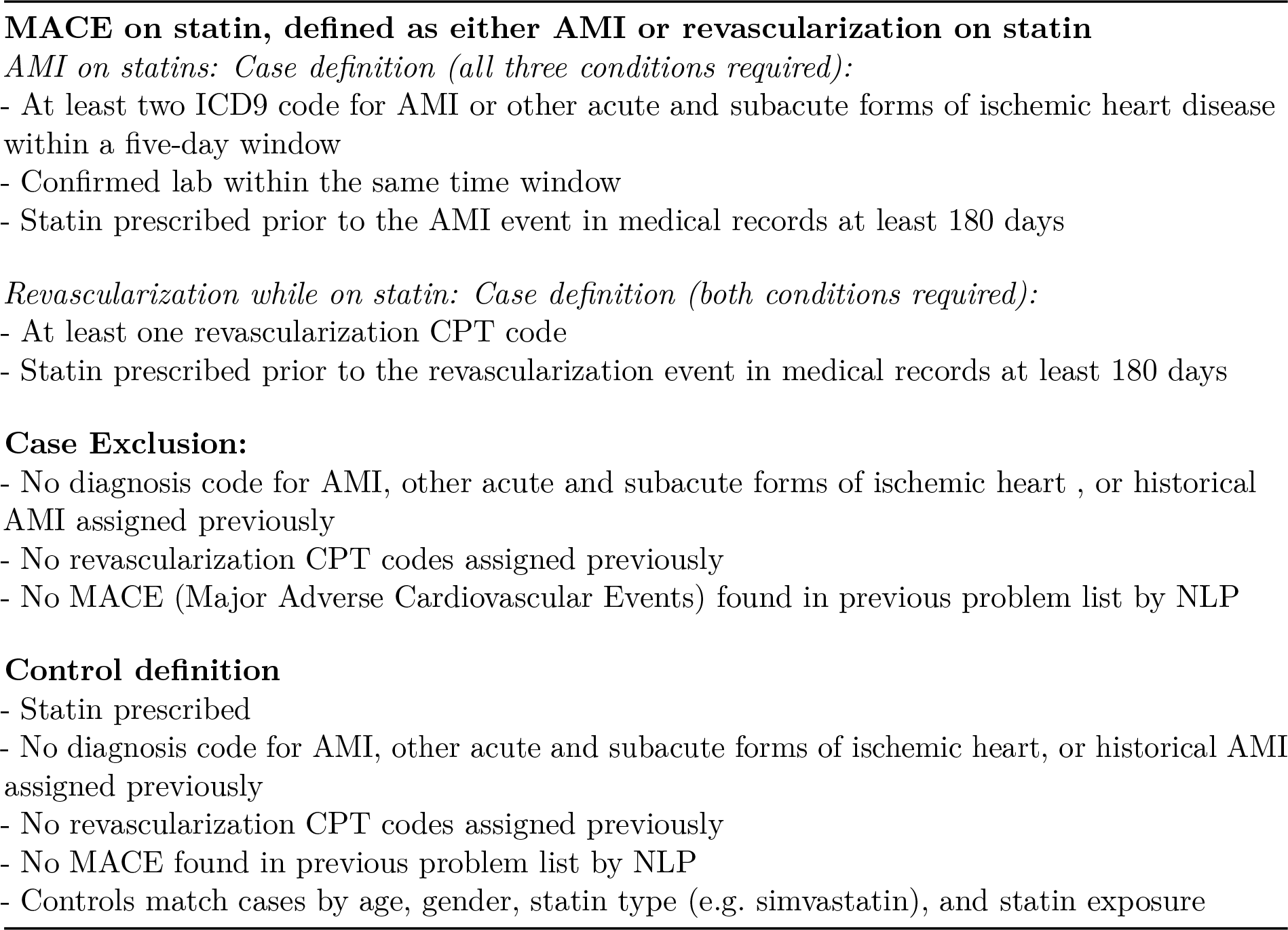
Inclusion and Exclusion Criteria

### Data Pre-Processing

Data obtained from dbGaP were in Plink format. The XY pseudo-autosomal region was recoded, and then the resulting file was converted to a multi-sample VCF file. The VCF file chromosome and positions were recoded based on the Illumina variant IDs from the HumanOmniExpressExome manifest file Infinium OmniExpressExome-8 v1.6 for GRCh38. Positions with ambiguous chromosome or positions were filtered from the resulting VCF file. Finally, filters were applied so that variants included in the final analysis were autosomal variants with a minor allele frequency of at least one percent. A PLINK format phenotype file was created from the original phenotype file from dbGaP. This created the necessary ID columns and selects the phenotype column corresponding to MACE. The resulting VCF file was converted to the transposed PLINK (tped) format with PLINK and carried forward for additional analyses.

### Random Forest Iterative Feature Reduction and Selection (RF-IFRS)

PLINK format files were read into an R environment with the GenABEL package.^12^ To account for the sensitivity of random forest (RF) models to group imbalance, we weighted cases and controls so that the probability of selecting either from a bootstrapped population was equivalent. To provide a reference comparison, we performed genome-wide association (GWA) analysis on the data with Plink version 1.9 using an additive model with no covariates.^13^ We used a two-step process for feature selection that sought to overcome computational limits of analyzing highly dimensional GWAS data. The first stage of feature reduction was performed using the Ranger package for R, in which the forest was grown with a mtry fraction of 1/3, and 1000 trees.^14^ Considering that this method incorporates the full breadth of data, we used the corrected impurity score implemented by Nembrini and colleagues, which overcomes the sensitivity of GINI importance to allele frequency while allowing a practical computing time compared to the more robust permutation score.^15^ This method is computationally fast, but relatively non-specific and produces false-positives similar to that of a traditional GWAS.

Features with p values of < 0.01 were selected for secondary feature selection with r2VIM, which incorporates multiple RF models to build a consensus permutation importance.^16^ It was re-implemented by Degenhardt and colleagues to support the ranger package, which allows for parallel tree building and much faster execution in the Pomona package.^17^ For our implementation, we cloned the Pomona repository and modified it so that it would accept input from a GenABEL object. The resulting custom r2VIM implementation was run with 11 sequentially grown RF models with 10,000 trees per forest using, an mtry fraction of 1/3, and nodes were limited to a maximum of 10% of the total population to limit tree depth. Features from the first forests with a minimum permutation importance of at least one in each forest were selected for estimation of association and interaction. The final (11th) forest was saved for the ensemble-method for epistasis selection.

### Testing for Epistasis

The ensemble method for epistasis estimation was implemented based on the work by Schmalohr and colleagues.^18^ We implemented methods for testing paired selection frequency (i.e. the probability that a variable will be included in the same decision tree) and selection asymmetry (i.e. the probability of a variant favoring a particular node when following another variant).^18^ These methods provides the means to detect AND and XOR epistasis. To create a final estimate for the presence of an interaction, p values from each method were combined using the Fisher method.^19^

Variant-pair p values were adjusted with the Benjamini-Hochberg FDR method, and pairs with FDR of less than 0.05 were retained for further analysis.^20^ Selected variant pairs were converted to dummy variables, and all pair-wise genotype permutations were compared with logistic regression. The minimum pairwise interaction p value was retained for each variant pair. Interaction p values were adjusted with the Benjamini-Hochberg FDR method.

### Poly-Epistatic Risk and Pathway Analysis

To extend beyond pair-wise interactions, pairwise interacting variants were condensed based on overlap. For example, A|B and A|C = A|B|C. Decision trees were built from the resulting variant interaction networks to visualize relationships and odds ratios based on multiple variants. Decision trees were built with the ctree function in the Party package for R.^21^ Odds ratios for terminal nodes were normalized to the overall odds of being a case.

To incorporate basic mechanistic insight, data were analyzed through the use of Ingenuity® Variant Analysis(tm) software (https://www.qiagenbioinformatics.com/products/ingenuity-variant-analysis) from QIAGEN, Inc. Top diseases and bio-functions relevant to MACE and CVD were reported with correlation to identified decision trees, then filtered for at least two genes involved and a FDR corrected p value of less than 0.05.

## Results

### Demographics

Demographic data are summarized in **Table <u>2</u>**. Our analysis incorporated genetic variant data and sex, which were available for all subjects. Random forest models do not tolerate missing values and require either imputation or exclusion to include variables with missing data. Given the focus on epistasis in this analysis, non-genetic variables were only included if they were defined in all cases and controls. Weight, and height were frequently missing in controls, and were therefore not used. Age was only available as “age of first event,” which limited its utility in comparing cases and controls. More than 99% of subjects in this population are reported as white, so no correction for race was incorporated. Therefore, only sex was incorporated into models. Sex is also a well-established predictor of risk for MACE.

**Table 2:** Population Demographics

| Variable | Case | Control | p |
| --- | --- | --- | --- |
| Count | 1735 | 4155 | - |
| Female(%) | 31.2% | 38.0% | <0.001 |
| White(%) | 99.4% | 99.3% | 0.507 |
| BMI(Mean±SD) | 28.57±7.035 | 29.03±7.37 | 0.253 |
| Age First MACE(Median±IQR) | 65±16 | N/A | 1 |

### Feature Selection with RF-IFRS

After pruning, there were 637,732 variants and 5,890 subjects in the cohort. Of the subjects, there were 1,735 cases and 4,155 controls. Evaluation of additive statistical association did not identify any variants that met genome-wide significance (5*10^−8^). The RF with the corrected impurity importance measure identified 6,688 variants with a corrected-impurity p value less than 0.01. As with statistical association, no variants met genome-wide significance. The 6,688 initially selected variants were extracted from the full dataset and analyzed with r2VIM. This identified 49 genetic variants in addition to sex with a minimum permutation importance value of at least one. Results from these analyses are shown in **Figure <u>1</u>**.

**Figure 1:**
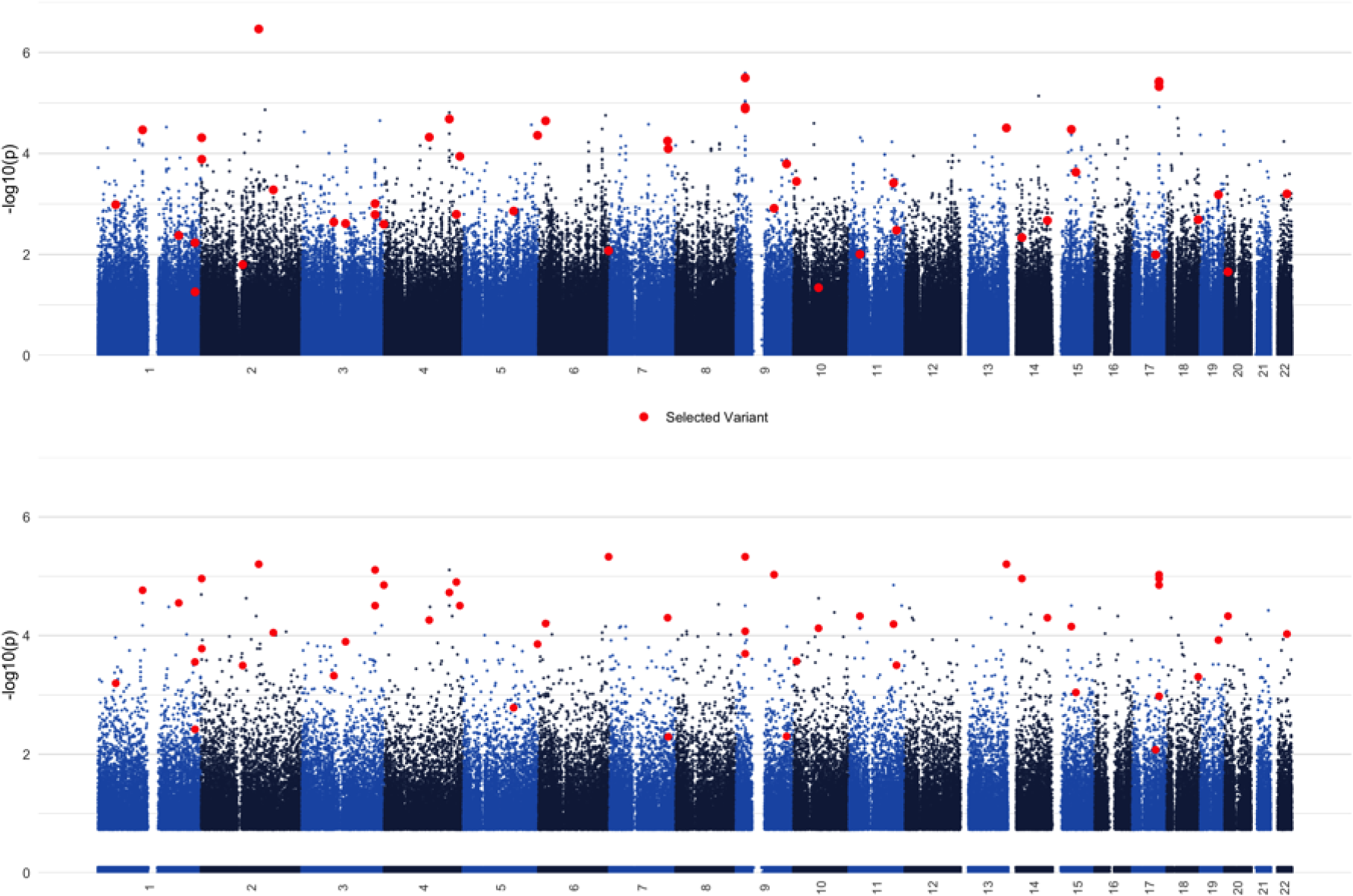
Manhattan plots for statistical GWAS analysis with PLINK (top) vs. the initial RF model with ranger (bottom). Red dots correspond to variants that were selected with r2VIM, and show that a purely statistical approach fails to identify variants that are likely relevant to the outcome due to interactions.

### Epistasis Screening

Paired selection frequency results identify variant-pairs that co-occur in decision trees more often, as often, or less often than predicted based on individual variant selection. Variants that are selected together more often than expected suggests a greater phenotype prediction from both variants together, and selection less often than expected suggests that co-occurence comes at a cost to phenotype prediction (i.e. variants are correlated and/or in linkage disequilibrium). **Figure <u>2</u>** shows the distribution of expected tree co-occurence for each variant pair. Using an alternative hypothesis of “greater than expected” in a binomial test allows sensitive selection of variant pairs that are chosen more often than predicted (red). We found evidence of epistasis in 16 variant-variant pairs **Table <u>3</u>**. Additionally, five variants showed significant interaction with sex.

**Figure 2:**
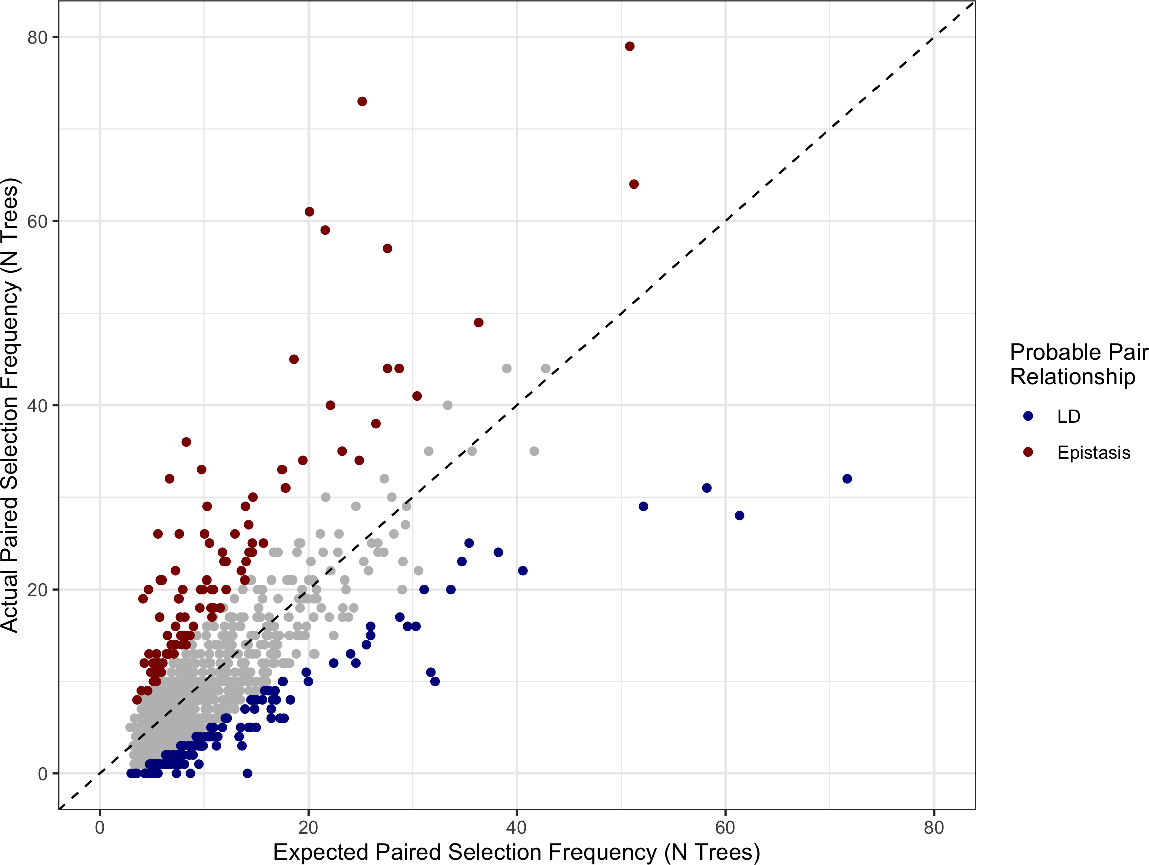
Paired selection frequency based on the combined independent variant probabilities (X axis) vs. the actual frequency of variants being selected together in a decision tree. Variants that are selected together at a lower-than-expected frequency are expected to be correlated with respect to the outcome, suggesting that they are in linkage disequilibrium (blue). Variants selected together more often than expected (red) are predicted to exhibit epistasis with respect to the phenotype.

**Table 3:**
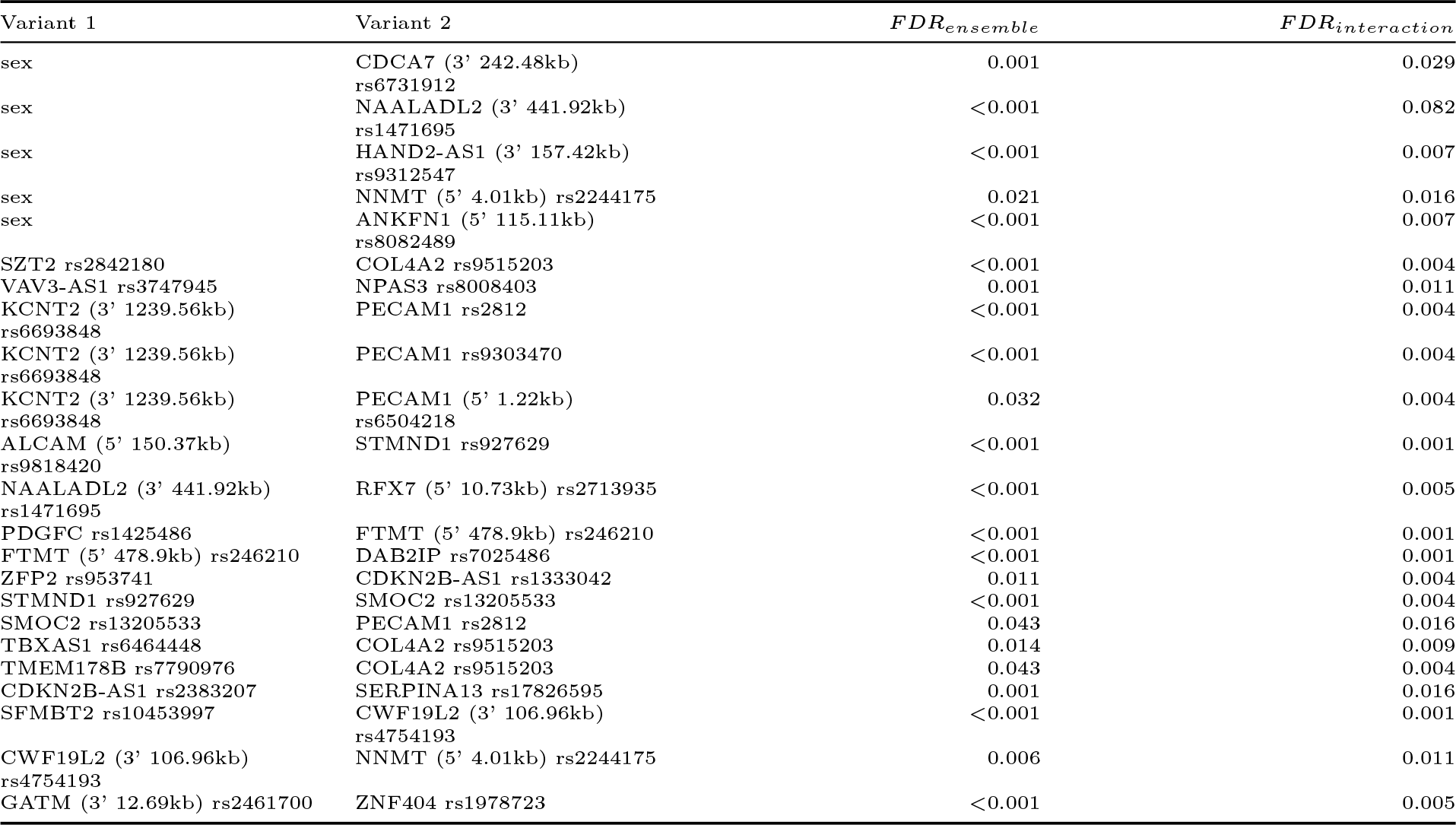
Significant Ensemble and Regression Variant Pairs

| Variant 1 | Variant 2 | $FDR_{ensemble}$ | $FDR_{interaction}$ |
| --- | --- | --- | --- |
| sex | CDCA7 (3' 242.48kb)<br>rs6731912 | 0.001 | 0.029 |
| sex | NAALADL2 (3' 441.92kb)<br>rs1471695 | <0.001 | 0.082 |
| sex | HAND2-AS1 (3' 157.42kb)<br>rs9312547 | <0.001 | 0.007 |
| sex | NNMT (5' 4.01kb) rs2244175 | 0.021 | 0.016 |
| sex | ANKFN1 (5' 115.11kb)<br>rs8082489 | <0.001 | 0.007 |
| SZT2 rs2842180 | COL4A2 rs9515203 | <0.001 | 0.004 |
| VAV3-AS1 rs3747945 | NPAS3 rs8008403 | 0.001 | 0.011 |
| KCNT2 (3' 1239.56kb)<br>rs6693848 | PECAM1 rs2812 | <0.001 | 0.004 |
| KCNT2 (3' 1239.56kb)<br>rs6693848 | PECAM1 rs9303470 | <0.001 | 0.004 |
| KCNT2 (3' 1239.56kb)<br>rs6693848 | PECAM1 (5' 1.22kb)<br>rs6504218 | 0.032 | 0.004 |
| ALCAM (5' 150.37kb)<br>rs9818420 | STMND1 rs927629 | <0.001 | 0.001 |
| NAALADL2 (3' 441.92kb)<br>rs1471695 | RFX7 (5' 10.73kb) rs2713935 | <0.001 | 0.005 |
| PDGFC rs1425486 | FTMT (5' 478.9kb) rs246210 | <0.001 | 0.001 |
| FTMT (5' 478.9kb) rs246210 | DAB2IP rs7025486 | <0.001 | 0.001 |
| ZFP2 rs953741 | CDKN2B-AS1 rs1333042 | 0.011 | 0.004 |
| STMND1 rs927629 | SMOC2 rs13205533 | <0.001 | 0.004 |
| SMOC2 rs13205533 | PECAM1 rs2812 | 0.043 | 0.016 |
| TBXAS1 rs6464448 | COL4A2 rs9515203 | 0.014 | 0.009 |
| TMEM178B rs7790976 | COL4A2 rs9515203 | 0.043 | 0.004 |
| CDKN2B-AS1 rs2383207 | SERPINA13 rs17826595 | 0.001 | 0.016 |
| SFMBT2 rs10453997 | CWF19L2 (3' 106.96kb)<br>rs4754193 | <0.001 | 0.001 |
| CWF19L2 (3' 106.96kb)<br>rs4754193 | NNMT (5' 4.01kb) rs2244175 | 0.006 | 0.011 |
| GATM (3' 12.69kb) rs2461700 | ZNF404 rs1978723 | <0.001 | 0.005 |

Condensing variant-pairs based on overlap resulted in six variant networks **Figure <u>3</u>**. We found networks that involved intergenic variants, for which the functional consequence is not clear. This is evident in network 1, where sex precedes four intergenic variants, most of which are more than 100kb away from the nearest gene. Gene-variant networks show diversity in odds for experiencing MACE, with individual node odds ratios reflecting the contribution of multiple variant effects through additive and non-additive relationships. We found that a small subset of subjects carrying *COL4A2* rs9515203 (T/T), *TMEM178B* rs7790976 (G/G), *SZT2* rs2842180 (C/T), and *TBXAS1* rs6464448 (G Allele) showed the highest increase in MACE risk (Network 2, OR = 4.53, p < 0.001). Variant effects analysis showed evidence of gene networks associated with angiogenesis, endothelial cell development and function, carotid artery disease, and development of vasculature (minimum FDR = 0.019) **Table <u>4</u>**.

**Figure 3:**
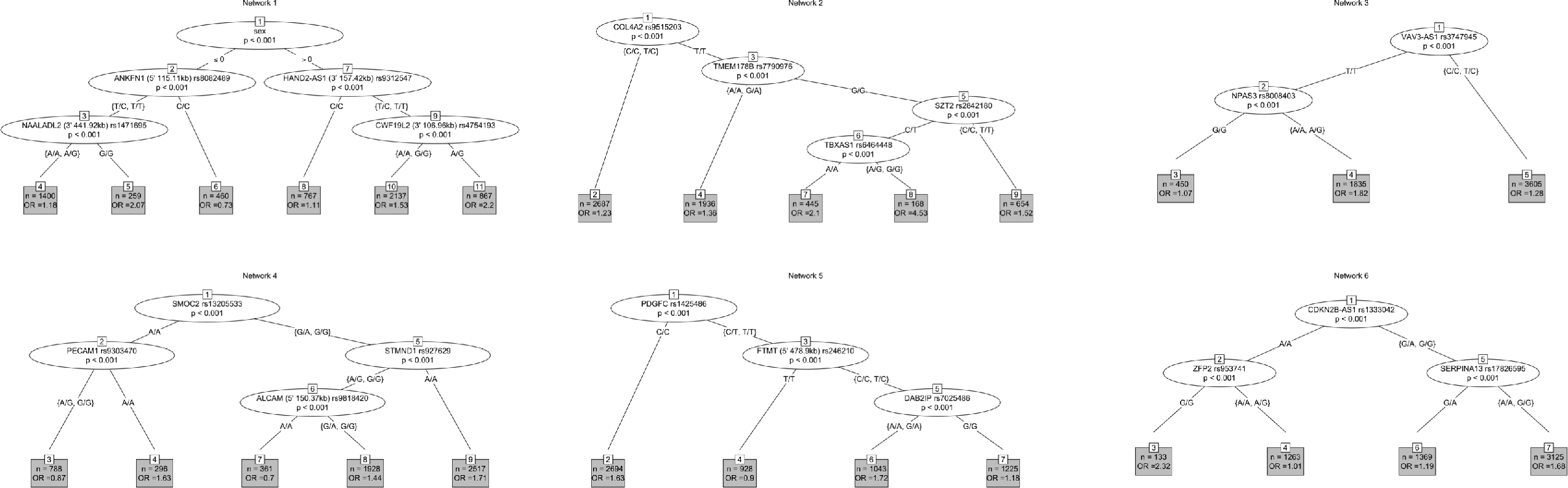
Decision trees incorporating overlapping epistasis-variant-pairs show six unique networks of genes and variants. Odds ratios in terminal nodes represent the odds of experiencing on-statin MACE in someone carrying the collection of alleles shown in the network relative to those who did not carry those variants. This shows a practical interpretation of epistasis findings that might be more practical to incorporate into clinical practice.

**Table 4:** Gene network associated disease processes

| Diseases or Functions | Genes | FDR |
| --- | --- | --- |
| Angiogenesis | *ALCAM CDKN2B COL4A2 DAB2IP*<br>*PDGFC PECAM1 SMOC2 VAV3* | 0.0188 |
| Carotid artery disease | *NNMT VAV3* | 0.034 |
| Development of vasculature | *ALCAM CDKN2B COL4A2 DAB2IP*<br>*NPAS3 PDGFC PECAM1 SMOC2 VAV3* | 0.0188 |
| Endothelial cell development | *COL4A2 PDGFC PECAM1 SMOC2* | 0.0291 |
| Formation of blood vessel | *CDKN2B COL4A2 PECAM1* | 0.0242 |
| Formation of endothelial tube | *COL4A2 PECAM1* | 0.0291 |
| Function of endothelial tissue | *PECAM1 VAV3* | 0.0188 |
| Migration of endothelial cells | *ALCAM COL4A2 PECAM1 SMOC2 VAV3* | 0.0188 |
| Quantity of endothelial cells | *ALCAM PDGFC* | 0.023 |
| Vasculogenesis | *ALCAM CDKN2B COL4A2 PDGFC*<br>*PECAM1 SMOC2* | 0.0242 |

## Discussion

### Risk Variants and Interactions for CVD

This study was a genome-wide study for variant-variant interactions (epistasis) associated with on-statin MACE. We found six variant networks that show a diverse range of genetic interactions that predict increased or decreased risk for on-statin MACE. Our findings show that RF-IFRS produces polygenic predictors of risk for on-statin MACE, suggesting that limitations of low effect-sizes can be overcome by studying variant networks to produce a final odds ratio.

### Angiogenesis, Endothelial Function, and Vasculogenesis in CVD

Angiogenesis refers to the formation of new capillary beds from existing vasculature, whereas vasculogenesis refers to the formation of *de novo* vascular networks (i.e. during embryonic development).^22^ Among others, *ALCAM, CDKN2B, COL4A2, DAB2IP, PECAM1, SMOC2, VAV3*, and *PDGFC* are related to either angiogenesis and/or vasculogenesis. These genes were included in five out of six networks that we identified, suggesting that these processes are relevant to risk for MACE and on-statin MACE.

### RF-IFRS replicates existing gene associations with CVD and incorporates novel interactions

Network one incorporated interactions with sex and variants in four intergenic regions. These variants flanked the nearest genes (*NAALADL2, HAND2-AS1, NNMT*, and *ANKFN1*) by up to 450kb. Drawing mechanistic insight from these interactions is not practical or necessarily advisable without further mechanistic analysis. However, this finding suggests that the association of male sex with higher risk for on-statin MACE is connected to diverse genetic components that might connect to chromatin structure, un-annotated regulatory RNA genes (e.g. lncRNA, Micro-RNA, etc).

Network two shows a relationship between *COL4A2* rs9515203, *TMEM178B* rs7790976, *SZT2* rs2842184, and *TBXAS1* rs6464448. *COL4A2* (Collagen Type IV Alpha 2 Chain) codes for the collagen IV peptide *α* 2 chain, which is a component of the basement membrane surrounding the endothelium of blood vessels.^23^ *COL4A2* rs9515203 has previously shown association with sub-clinical atherosclerosis^24^, and coronary artery disease.^25,26^ Other variants in *COL4A2* and *COL4A1* show associations with risk for MI, atheroslcerotic plaque stability, and vascular stability.^23^ The role of *SZT2* (Seizure threshold 2 homolog) rs2842184 in CVD is not clear, and may not indicate a direct mechanism. A recent proteomic study of plasma protein expression in patients with CVD found decreased plasma levels of SZT2 in patients with CVD. The authors suggested that this might be connected to increased mTORC1 signalling in patients with CVD, but this mechanism has not been tested.^27^ *TMEM178B* (Transmembrane Protein 178B) codes for a transmembrane protein that is highly expressed in cardiac tissue, among others. The role of the rs7790976 variant is not clear in this network. *TBXAS1* (Thromboxane A synthase 1) codes for Thromboxane A Synthase 1, which is expressed in several tissues including platelets. Thromboxane is a potent vasoconstrictor that causes vasoconstriction and platelet aggregation. The rs6464448 variant has not been previously associated with a phenotype, and the role of genetic variation connecting *TBXAS1* to CVD outcomes is not clear. However, *TBXAS1* has been recently proposed as a potential drug target for CVD.^28^

Network three is comprised of an interaction between *VAV3-AS1* rs3747945 and *NPAS3* rs8008403. *VAV3* (Vav Guanine Nucleotide Exchange Factor 3) is important to the migration of smooth muscle cells, which suggests that it has a role in vascular proliferation.^29^ *VAV3-AS1* is an RNA gene coding for anti-sense *VAV3*, which might regulate expression of *VAV3*.^30^ *VAV3-AS1* rs3747945 has not been previously associated with cardiovascular disease related outcomes, but further supports the role for vasculogenesis in risk for MACE. *NPAS3* (Neuronal PAS Domain Protein 3) rs8008403 has not been previously associated with cardiovascular disease related outcomes, but another variant in *NPAS3* (rs17460823) was associated with C-reactive protein in patients taking fenofibrate.^31^ The mechanistic connection of these variants/genes is difficult to determine, but might be linked to development of gross anatomy of the cardiovascular system, or to remodeling associated with CVD.

Network four connects *SMOC2* rs13205533, *PECAM1* rs9303470, *STMND1* rs927629, and a variant (rs9818420) approximately 150kb upstream from *ALCAM*. This network appears to be related to vascular homeostasis and proliferation. *SMOC2* (SPARC-related modular calcium-binding protein 2) modulates calcium homeostasis, and might be relevant to blood vessel calcification.^32^ *SMOC2* rs13205533 has not been previously associated with cardiovascular disease related outcomes. *PECAM1* (Platelet And Endothelial Cell Adhesion Molecule 1) is important for the maintenance of vascular endothelial integrity, and endothelial cells that express PECAM1 are more resilient to the inflammatory response from vascular barrier damage.^33,34^ *PECAM1* rs9303470 has not been previously associated with cardiovascular disease related outcomes, but other variants in *PECAM1* have been found to be associated with CAD.^33^ *PECAM1* shares similar function with *ALCAM* (Activated leukocyte cell adhesion molecule), and both seem to play roles in CVD.^35^ Higher levels of the ALCAM protein have been associated with poor CV outcomes including CV death in patients presenting with ACS.^35^ *STMND1* (Stathmin Domain Containing 1) Variants in *STMND1* have been associated with stroke in African Americans, though rs927629 has not been previously reported with CVD.^36^

Network five shows interactions between genes relevent to angiogenesis, including *PDGFC* rs1425486 and *DAB2IP* rs7025486. Variants in *PDGFC* (Platelet Derived Growth Factor C) and other *PDGF* genes have been associated with angiogenesis and CVD.^37^ PDGFC likely promotes angiogenesis independently of VEGF, which might support a role in CVD development and/or vascular remodeling.^38^ *PDGFC* rs1425486 has not been previously associated with cardiovascular disease related outcomes. *DAB2IP* (DAB2-interacting protein) is expressed widely in the cardiovascular system and it is believed to be an inhibitor of VEGF-2 signalling and thus an inhibitor of angiogenesis.^39^ Multiple variants in *DAB2IP* have been associated with CAD,^40^ and rs7025486 is associated with abdominal aortic aneurysm.^41^

Network six includes interactions between *CDKN2B-AS1* rs1333042, *ZFP2* rs953741, and *SERPINA13* rs17826595. *CDKN2B-AS1* (cyclin-dependent kinase inhibitor 2B antisense RNA 1) is an RNA gene that regulates the expression of *CDKN2B*. CDKN2B is an inhibitor of cellular proliferation, though its direct role in CVD is not clear. Numerous variants in *CDKN2B-AS1*, including rs1333042, have been associated with CHD.^42^ *ZFP2* (Zinc Finger Protein) is a regulator protein. Variants in *ZFP2* are associated with MI in African Americans,^36^ though *ZFP2* rs953741 has not been previously associated with cardiovascular disease related outcomes. *SERPINA13* (Serpin Family A Member 13) is a pseudogene, and it is not clear what its role is in CVD. *SERPINA13* rs17826595 has not been previously associated with cardiovascular disease related outcomes. Other members of the *SERPIN* gene superfamily are related to cardiovascular system development and regulation.^43^

### Limitations

The RF-IFRS method is a novel approach to genome-wide epistasis that incorporates statistics and interpretable ML methods. The definition of MACE used in this study is less broad than is commonly used in the CVD literature. Notably, ischemic stroke and CV death are not included in the definitions, which is relevant to the generalizability of these findings to other studies that evaluate MACE as an outcome. This study was carried out in a single cohort of patients without replication, however, the RF procedure performs thousands of random samples from the dataset to determing feature importance. While this is not as robust as independent replication, it might help mitigate the bias associated with genetic association studies carried out in a single cohort. We did not split the cohort into training and testing groups or perform hyperparameter tuning, which are often done when developing a predictive ML model. However, the objective was not to generate a highly predictive ML model, but rather to use the organic structure of the RF approach to identify important variants and interactions. This is also relevant to statistical power, and given that this study found no significant variants with traditional GWAS we opted to keep the entire cohort together to maximize power. Due to the RF inclination to discover LD organically, we did not perform LD pruning. We also did not perform imputation to limit the computational overhead required for the RF model training. This study does not include causal analysis of individual SNPs, thus we do not suggest that the reported variants are necessarily causal. Finally, we did not have access to more extensive clinical data. Further analysis and replication ought to evaluate if findings correspond to degree of lipid control.

## Conclusions

A RF driven method for feature reduction and selection applied to a GWAS-scale dataset identified six epistasis-networks that may provide insight into the risk for on-statin MACE. This method also provides interpretable results, which may produce a more physiologically relevant assessment of odds and risk for an outcome than PRS. We found that variants related to angiogenesis and vasculogenesis are associated with odds of on-statin MACE. These findings present a unique opportunity for the incorporation of multiple low-effect size variants in the prediction of drug success in preventing CVD events.

## Data Availability

Controlled access data from dbGaP cannot be shared. Summary data, intermediate outputs, and methods are available in GitHub.

https://github.com/sadams-lab/manuscript_onstatin-mace-GWES

## Acknowledgements

The dataset used for this study were obtained from Vanderbilt University Medical Center’s BioVU, which is maintained in the NCBI Database for Genotypes and Phenotypes dbGaP.

